# Circulating Plasma Biomarkers in Biopsy-Confirmed Kidney Disease: Results from the Boston Kidney Biopsy Cohort

**DOI:** 10.1101/2021.04.14.21255472

**Authors:** Insa M. Schmidt, Suraj Sarvode Mothi, Parker C. Wilson, Ragnar Palsson, Anand Srivastava, Zoe A. Kibbelaar, Min Zhuo, Afolarin Amodu, Isaac E. Stillman, Helmut G. Rennke, Benjamin D. Humphreys, Sushrut S. Waikar

## Abstract

**Background:** Biomarkers for non-invasive assessment of histopathology and prognosis are needed in patients with kidney disease.

**Methods:** Using a proteomics assay, we measured a multi-marker panel of 225 circulating plasma proteins in a prospective cohort study of 549 individuals with biopsy-confirmed kidney diseases and semi-quantitative assessment of histopathology. We tested the associations of each biomarker with histopathologic lesions and the risks of kidney disease progression (defined as ≥40% decline in eGFR or ESKD) and death.

**Results:** After multivariable adjustment and correction for multiple testing, 46 proteins associated with different histopathologic lesions. The top performing markers positively associated with acute tubular injury and interstitial fibrosis and tubular atrophy were kidney injury molecule-1 (KIM-1) and V-set and immunoglobulin domain-containing protein 2 (VSIG2). 30 proteins were significantly associated with kidney disease progression and 35 with death. The top performing markers for kidney disease progression were placental growth factor (PGF; HR 5.4, 95% CI 3.4 to 8.7) and BMP and Activin Membrane Bound Inhibitor (BAMBI; HR 3.0, 95% CI 2.1 to 4.2); the top performing markers for death were TRAIL-receptor-2 (TRAIL-R2; HR 2.9, 95% CI 2.0 to 4.0) and CUB Domain Containing Protein-1 (CDCP1; HR 2.4, 95% CI 1.8, 3.3).

**Conclusion:** We identified several biomarkers associated with kidney disease histopathology and prognosis – many of which have not been reported previously and may represent important avenues for future research.

## Introduction

The two measures used to diagnose and stage kidney diseases, albuminuria and the estimated glomerular filtration rate (eGFR), do not provide specificity about kidney histopathological lesions and are imperfect prognostic markers. Novel biomarkers with greater specificity for histopathologic lesions will enhance clinical phenotyping of kidney diseases and may provide an important milestone toward precision medicine.

Several biomarkers of kidney inflammation and tubular injury have been shown to prognosticate the risk of adverse clinical outcomes in patients with kidney diseases,^1-4^ but these biomarkers only represent a small segment of the human blood proteome. Proteomics-based approaches add to this by allowing for an unbiased protein discovery that does not entirely depend on pre-existing pathophysiologic knowledge. The history of biomarker studies in other clinical disciplines illustrates how large-scale proteomics approaches can identify important protein markers for improved risk prediction^5^ and the development of new therapeutic targets.^6^ Fewer studies, however, have made use of these techniques for investigation of human kidney disease.^7-9^

In this study, we used a novel proteomics assay to measure a multi-marker panel of 225 plasma proteins in a prospective cohort study of individuals with biopsy-confirmed kidney diseases and adjudicated semi-quantitative assessment of histopathology. We tested the associations of each biomarker with kidney histopathologic lesions and the risks of subsequent kidney disease progression and death, and explored pathways enriched in the plasma proteome of patients with kidney diseases using the Reactome pathway database.

## Methods

### Study Population

The Boston Kidney Biopsy Cohort (BKBC) is a prospective, observational cohort study of patients undergoing native kidney biopsy at three tertiary care hospitals in Boston, Massachusetts, including Brigham and Women’s Hospital, Massachusetts General Hospital, and Beth Israel Deaconess Medical Center. Details of the study design have been previously described.^10^ The study includes adults ≥18 years of age who underwent a clinically indicated native kidney biopsy between September 2006 and October 2018. Exclusion criteria were the inability to provide written consent, severe anemia, pregnancy, and enrollment in competing studies. Patients provided blood and urine samples on the day of kidney biopsy. For this study, we evaluated 549 participants with available plasma samples that included both common and more rare forms of kidney diseases. The Partners Human Research Committee (the Brigham and Women’s Hospital Institutional Review Board) approved the study protocol which is in accordance with the principles of the Declaration of Helsinki.

### Sample Collection, Proteomics Assays, and Exposures

Blood samples were collected from study participants on the day of biopsy, aliquoted, and immediately stored at −80°C. Aliquots were analyzed at Olink using high-throughout, multiplex immunoassays^11^ on three commercially available panels named Inflammation, Organ Damage, and Cardiovascular II. Each panel consists of 92 biomarker proteins that were chosen based on their potential relevance in various pathological processes. All protein values are expressed as normalized protein expression (NPX) values on a log_2_ scale. We included 5% blind split replicates in addition to BKBC samples. Of the 276 proteins included in the three panels, we used 225 biomarkers as the primary exposures for statistical analyses that were non-overlapping across the panels and passed the following quality control metrics: Coefficients of variation (CV) <10% from blind split replicates; standard deviation (SD) of internal Olink controls <0.2, and incubation or detection control which deviated < +/- 0.3 from the median value of all samples on the plate. As an additional quality control, we included multiple plasma aliquots from two patients, one with high and one with low eGFR, which were spread randomly in dummy labeled tubes across the shipment boxes. The mean CVs were 5.5 (+/- 4.3) and 4.9 (+/- 4.6), respectively, for the 225 biomarkers.

### Histopathologic Outcomes

Kidney biopsy specimens were adjudicated under light microscopy by two experienced kidney pathologists who provided semiquantitative scores of kidney inflammation, fibrosis, vascular sclerosis, and acute tubular injury (**Supplemental Table 1**). Methods to evaluate and score histopathologic lesions were previously described in detail.^10^ Of the 13 histopathologic lesions adjudicated, all were scored during study sessions except for grades of global or segmental glomerulosclerosis, which were taken from the biopsy report, because they were each calculated as a percentage of the total number of glomeruli. We combined endocapillary glomerular inflammation, extracapillary cellular crescents, focal glomerular necrosis, and fibrocellular crescents into a single dichotomous variable named ‘glomerular inflammation’ due to the relatively low prevalence and limited range of severity for each of those lesions in this cohort. All participants’ charts were reviewed alongside histopathologic evaluations to provide the final primary clinicopathologic diagnosis.

### Clinical Outcomes

The primary outcome was kidney disease progression, defined as ≥40% decline in eGFR or ESKD (initiation of dialysis or kidney transplantation). The secondary outcome was death. Data on eGFR during follow-up was obtained from the electronic medical record (EMR) and ESKD status was confirmed by reviewing the EMR and linkage with the US Renal Data System database. Mortality status was confirmed with the Social Security Death Index. Participants were followed until the occurrence of death, voluntary study withdrawal, loss to follow-up, or February 1, 2020.

### Covariates

Detailed patient information was collected at the biopsy visit, including demographics, medical history, medication lists, and pertinent laboratory data and stored using REDCap electronic data capture tools hosted at Partners Health Care. We obtained serum creatinine (SCr) from the EMR on the day of biopsy. In participants for whom this was unavailable, we measured SCr in available blood samples collected on the day of biopsy. We obtained spot urine protein-to-creatinine ratio or urine albumin-to-creatinine ratio from the date of kidney biopsy up to 3 months before biopsy from the EMR. SCr and urine creatinine were measured using a Jaffe-based method and urine albumin was measured by an immunoturbidometric method. The creatinine-based Chronic Kidney Disease Epidemiology Collaboration equation was used to calculate the eGFR.^12^

### Pathway Analyses

Pathway analyses were performed using Reactome.^13^ Reactome employs the Benjamini-Hochberg approach to provide a False Discovery Rate (FDR) that accounts for the number of tests performed. Statistical significance was assessed by permuting group labels using 1,000 permutations and pathways were ranked according to their p-values and FDR.

### Statistical Analysis

We summarized descriptive statistics as count with percentages for categorical variables and mean ± standard deviation or median with interquartile range for continuous variables. For skewed data distributions, we performed natural logarithmic transformation as appropriate. Unadjusted and adjusted multivariable linear regression models were used to assess associations of each plasma biomarker protein with histopathologic lesions. For these analyses, histopathologic lesions were dichotomized as less severe versus more severe lesions. We limited statistical analyses on histopathologic lesions to participants with adjudicated histopathology by both kidney pathologists (n=411, 75%) except for analyses of global or segmental glomerulosclerosis since they were taken from the biopsy report (n=549).

We performed time-to-event analyses to examine the associations of biomarkers with kidney disease progression and death. Cox proportional hazard models were stratified by site and adjusted for age, race, sex, log(proteinuria), eGFR, and primary clinicopathologic diagnostic category of kidney disease. For the outcome of kidney disease progression, we treated the data as interval censored because the exact date of the event may not be known. We evaluated the association between plasma biomarkers and subsequent kidney disease progression using a nonparametric survival function for interval-censored data.^14^ We confirmed no violations of the proportional hazards assumption through assessment of Schoenfeld residuals and used complete case analysis for the analyses as there were less than 5% missing data. To counteract the problem of multiple testing, we used Bonferroni correction (adjusted p-value = k × p_naive_, where k is the number of biomarkers included in the analyses (n=225) and p_naive_=0.05) for all reported p-values. Statistical analyses were performed using R Version 3.6.1 (R Foundation for Statistical Computing, Vienna, Austria).

## Results

### Baseline Characteristics

Baseline characteristics of the study cohort are shown in **Table 1**. The mean age was 51.6±16.7 years and 52.5% were women. The mean eGFR was 57.8±36.4 ml/min/1.73m^2^ and the median proteinuria (IQR) was 1.7 (0.4, 3.8) g/g creatinine. The most common primary clinicopathologic diagnoses were glomerulopathies (47.0%), diabetic nephropathy (11.7%), advanced glomerulosclerosis (11.3%), vascular disease (9.5%), and tubulointerstitial disease (8.6%).

**Table 1.** Baseline characteristics of the study cohort.

| **Baseline characteristics** | **n = 549** |
| --- | --- |
| **Clinical Characteristics** |  |
| Age, years | 51.6 ± 16.7 |
| Female | 288 (52.5) |
| **Race** |  |
| Black | 105 (19.1) |
| White | 347 (63.2) |
| Other | 97 (17.7) |
| eGFR, ml/min/1.73m<sup>2</sup> | 57.8 ± 36.4 |
| **CKD stage** |  |
| Stage 1 (eGFR > 90 ml/min/1.73m<sup>2</sup>) | 132 (24.0) |
| Stage 2 (eGFR 60-89 ml/min/1.73m<sup>2</sup>) | 92 (16.8) |
| Stage 3A (eGFR 45-59 ml/min/1.73m<sup>2</sup>) | 81 (14.8) |
| Stage 3B (eGFR 30-44 ml/min/1.73m<sup>2</sup>) | 91 (16.6) |
| Stage 4 (eGFR 15-29 ml/min/1.73m<sup>2</sup>) | 101 (18.4) |
| Stage 5 (eGFR < 15 ml/min/1.73m<sup>2</sup>) | 52 (9.5) |
| **Proteinuria, g/g creatinine** | 1.7 [0.4, 3.8] |
| **Reason for Biopsy\*** |  |
| Proteinuria | 312 (56.8) |
| Hematuria | 134 (24.4) |
| Nephrotic Syndrome | 69 (12.6) |
| Nephritic Syndrome | 14 (2.6) |
| Abnormal eGFR/Other | 336 (61.2) |
| **Primary Clinicopathologic Diagnosis** |  |
| Proliferative glomerulonephritis | 162 (29.5) |
| Non-proliferative glomerulopathy | 96 (17.5) |
| Diabetic nephropathy | 64 (11.7) |
| Advanced glomerulosclerosis | 62 (11.3) |
| Vascular disease | 52 (9.5) |
| Tubulointerstitial disease | 47 (8.6) |
| Paraprotein-related disease | 35 (6.4) |
| Other | 31 (5.6) |
| **Co-morbid Conditions** |  |
| Diabetes Mellitus | 122 (22.2) |
| Hypertension | 287 (52.3) |
| Systemic Lupus Erythematosus | 90 (16.4) |
| Hepatitis C | 11 (2.0) |
| Hepatitis B | 4 (0.7) |
| HIV | 6 (1.1) |
| Malignancy | 80 (14.6) |
| **Medications** |  |
| ACEi/ARB | 257 (46.8) |
| MRA | 14 (2.6) |
| Calcium channel blockers | 137 (25.0) |
| Beta-blockers | 166 (30.2) |
| Immunosuppression | 102 (18.6) |
| Corticosteroids | 123 (22.4) |
| **Clinical site** |  |
| Site 1 | 384 (69.9) |
| Site 2 | 104 (18.9) |
| Site 3 | 61 (11.1) |
| **Clinical endpoints** |  |
| ≥40% eGFR decline or ESKD | 170 (31.4) |
| Death | 82 (15.0) |

Data presented as mean ± standard deviation, median [interquartile range], and count with frequencies (%) for binary and categorical variables
\*Percentages do not add to 100 as there may have been more than 1 reason for a kidney biopsy SLE, Systemic Lupus Erythematosus; ACE, angiotensin converting enzyme inhibitor; ARB, angiotensin receptor blocker; MRA, mineralocorticoid receptor blocker; eGFR, estimated glomerular filtration rate; ESKD, end-stage kidney disease

### Plasma protein biomarkers associated with histopathologic lesions

Associations between plasma biomarkers and histopathologic lesions are shown in **Figure 1 and Supplemental Table 2**. After multivariable adjustment and correction for multiple testing, 46 plasma proteins were independently associated with different histopathologic lesions. By level of statistical significance, the top-performing biomarkers positively associated with more severe acute tubular injury (ATI) and interstitial fibrosis/tubular atrophy (IFTA) were kidney injury molecule-1 (KIM-1) and V-set and immunoglobulin domain-containing protein 2 (VSIG2), respectively (**Supplemental Table 2**). The top-performing biomarkers positively associated with other lesions included: Protein S100-A12 (EN-RAGE) with greater severity of glomerular inflammation; C-X-C motif chemokine 9 (CXCL9) with inflammation in the non-fibrosed interstitium; tissue factor (TF) with glomerular sclerosis, and tumor necrosis factor (TNF)-related apoptosis-inducing ligand (TRAIL) with mesangial expansion. Matrix metalloproteinase-7 (MMP-7) and fatty acid-binding protein 2 (FABP-2) were the top performing markers associated with more severe inflammation in the fibrosed interstitium and arteriolar sclerosis, respectively. Nine biomarkers were inversely associated with histopathologic lesions (**Supplemental Table 2**). Among those, stem cell factor (SCF) associated inversely with both acute tubular injury and inflammation in the non-fibrosed interstitium. There was no significant association between any of the biomarkers and segmental glomerulosclerosis or arterial sclerosis after multivariable adjustment.

**Figure 1.**
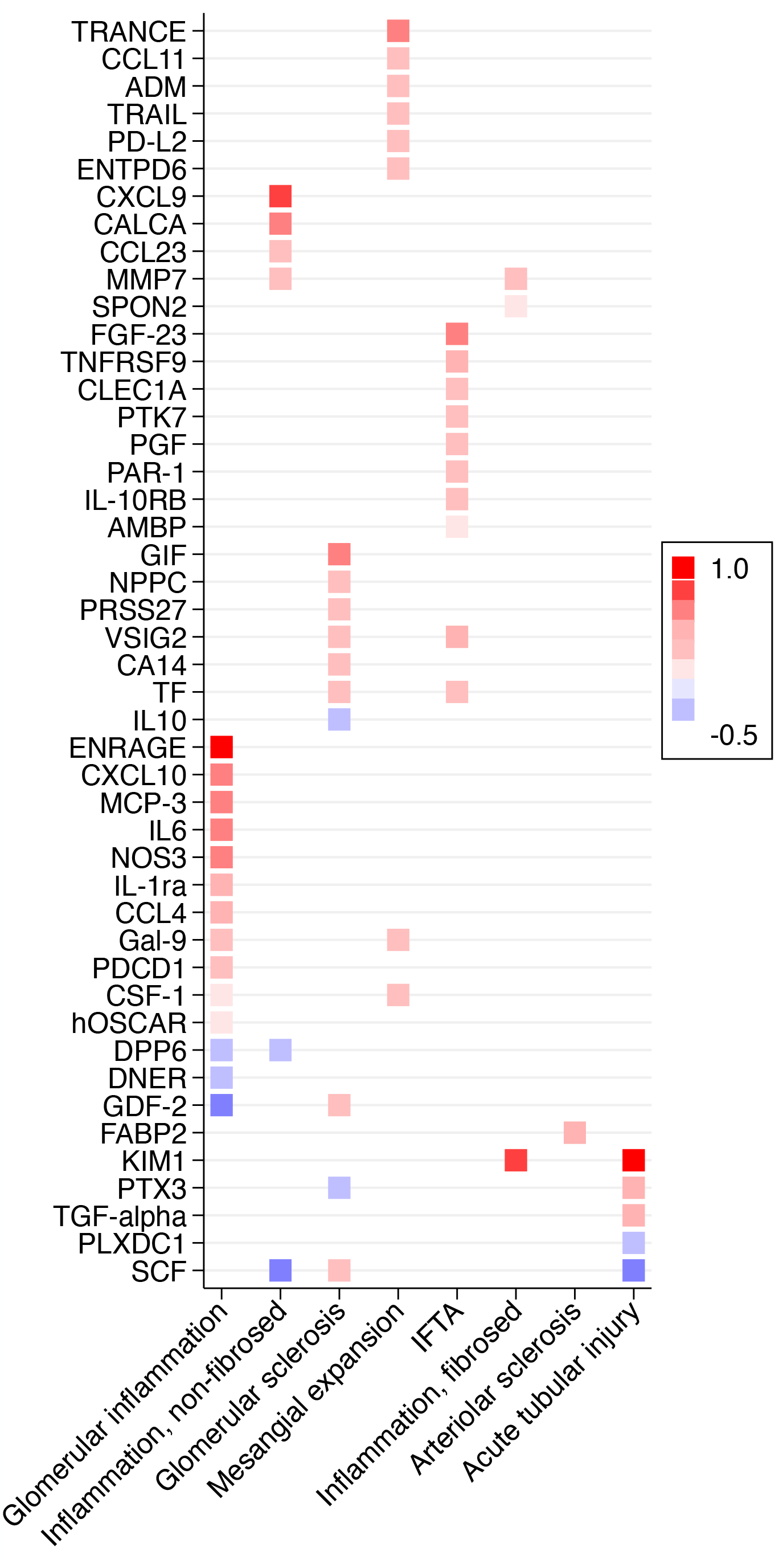
Circulating plasma proteins associated with histopathologic lesions in native kidney biopsy specimens. The heatmap shows associations between histopathologic lesions and biomarkers after Bonferroni correction. Beta coefficients are derived from multivariable linear regression models, adjusted for age, sex, race, and eGFR and displayed as colors ranging from blue to red. Reference is absence of lesion for glomerular inflammation and inflammation in the non-fibrosed interstitium; none/mild lesion severity for mesangial expansion, acute tubular injury, and arteriolar sclerosis; and 0-25% of cortical volume affected for global glomerulosclerosis, inflammation in the fibrosed interstitium, and interstitial fibrosis/tubular atrophy.

### Plasma protein biomarkers associated with kidney disease progression

170 individuals suffered kidney disease progression during a median follow-up of 51.8 months. Higher levels of 30 plasma proteins were independently associated with greater risk of kidney disease progression **(Figure 2A and Supplemental Table 3)**. In multivariable-adjusted models, higher levels of the following biomarkers were associated with greater risks of kidney disease progression (top 5 findings in the order of level of significance): Placental growth factor (PGF), BMP and activin membrane bound inhibitor (BAMBI), TNF-receptor superfamily-11A (TNFRSF-11A), TNF-related apoptosis-inducing ligand-R2 (TRAIL-R2/TNFRSF-10B), and C-X3-C motif chemokine ligand 1 (CX3CL1). Among all 30 proteins, several were interleukins (IL) and IL-receptors (IL-4RA, IL-10RB, IL-15RA, IL-16), hemostatic markers (thrombomodulin (TM), protease-activated receptor-1 (PAR-1), TF) and members of the TNF-superfamily (TNFRSF-9, TNFRSF-10A, TRAIL-R2/TNFRSF-10B, TNFRSF-11A). Cytokine signaling and activation of the clotting cascade were among the top ranked pathways associated with kidney disease progression (**Table 2**).

**Figure 2.**
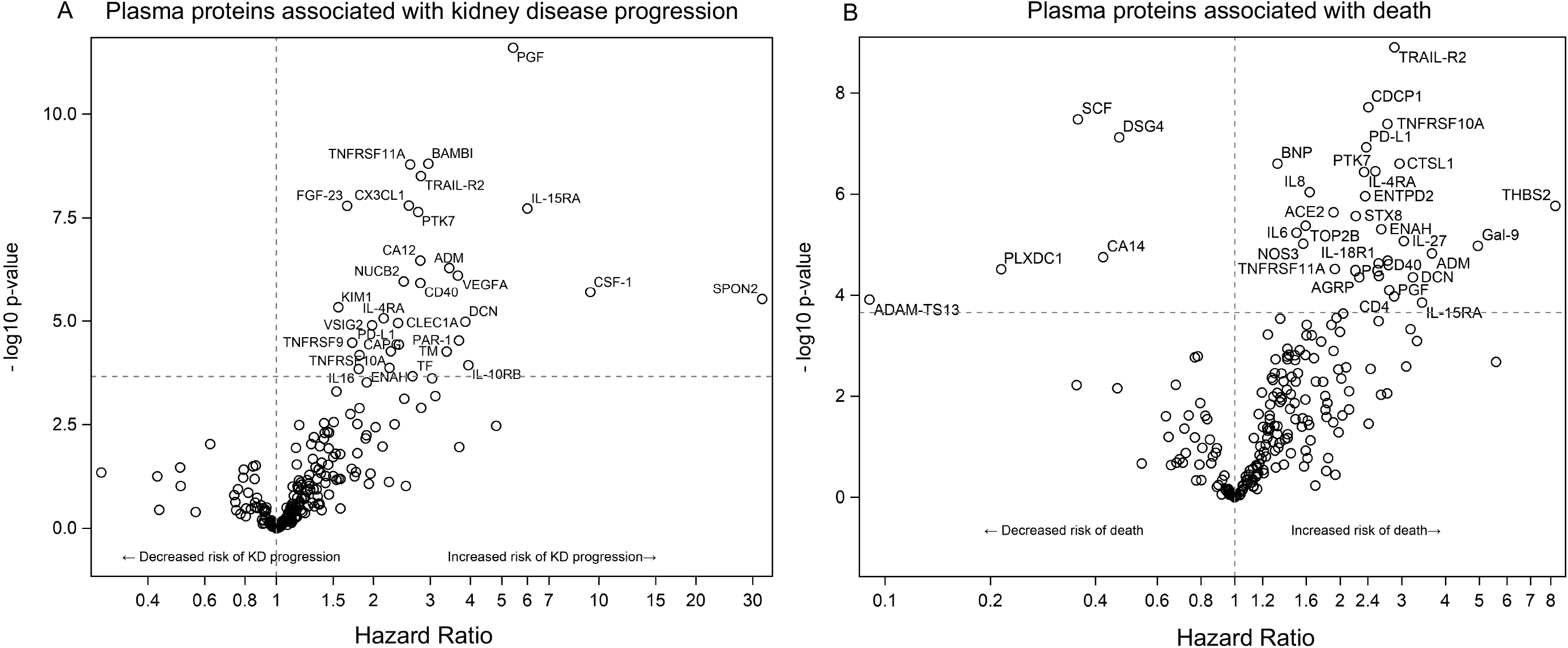
Circulating plasma proteins associated with kidney disease progression (A) and death (B). Results are derived from proportional hazards models adjusted for eGFR, race, age, sex, log(proteinuria), and primary clinicopathologic diagnosis. Horizontal dotted lines show Bonferroni-adjusted significance thresholds and vertical dotted lines mark the hazard ratio of 1. Name and abbreviation of each biomarker are shown in Supplemental Table 5. KD, kidney disease.

**Table 2.** Top-ranked pathways for plasma biomarkers associated with adverse clinical outcomes.

| **Top-ranked pathways for kidney disease progression** |  |  |  |  |
| --- | --- | --- | --- | --- |
| Rank | Biomarkers | Pathway* | p-value | FDR |
| 1 | IL-10RB, CSF-1, IL16 | Other Interleukin Signaling | 3.99e-05 | 0.007 |
| 2 | TF, TM, PAR-1 | Formation of Fibrin Clot (Clotting Cascade) | 1.67e-04 | 0.007 |
| 3 | TNFRSF9, VEGF-A, FGF-23, IL-4RA, IL-10RB, CD40, CSF-1, IL16, TNFRSF11A, IL-15RA | Cytokine Signaling in Immune system | 1.78e-04 | 0.007 |
| 4 | TRAIL-R2, TNFRSF10A | TRAIL signaling | 2.25e-04 | 0.007 |
| 5 | VEGF-A, PGF | VEGF ligand-receptor interactions | 2.25e-04 | 0.007 |
| **Top-ranked pathways for death** |  |  |  |  |
| Rank | Biomarkers | Pathway* | p-value | FDR |
| 1 | CD40-L, IL-4RA, CD4, IL-18R1, IL6, SCF, IL-15RA, IL-27, Gal-9, IL-8, TNFRSF11A, OPG | Cytokine Signaling in Immune system | 2.04e-05 | 0.004 |
| 2 | IL-27, IL-4RA, CD4, Gal-9, IL-18R1, IL-8, IL6, IL-15RA | Signaling by Interleukins | 5.77e-05 | 0.006 |
| 3 | TRAIL-R2, TNFRSF10A | TRAIL signaling | 2.88e-04 | 0.017 |
| 4 | CTSL1, ACE2 | Attachment and Entry | 4.48e-04 | 0.017 |
| 5 | TRAIL-R2, TNFRSF10A | Regulation by c-FLIP | 5.40e-04 | 0.017 |

### Plasma protein biomarkers associated with death

82 participants died during a median follow-up of 59.4 months, respectively. Higher levels of 30 proteins and lower levels of 5 proteins were independently associated with greater risk of death **(Figure 2B and Supplemental Table 4)**. The top 5 proteins positively associated with greater risk of death according to level of significance were TRAIL-R2/TNFRSF-10B, CUB domain containing protein 1 (CDCP1), TNFRSF10A, programmed death-ligand 1 (PD-L1/CD274), and brain natriuretic peptide (BNP). The 5 plasma proteins inversely associated with greater risk of death were SCF, Desmoglein-4 (DSG4), Carbonic anhydrase 14 (CA14), Plexin domain-containing protein 1 (PLXDC1), and disintegrin and metalloproteinase with thrombospondin motifs-13 (ADAMTS13). Among all proteins associated with death, there were several members of the TNF-superfamily (TNFRSF-9, TNFRSF-10A, TNFRSF-10B/TRAIL-R2, TNFRSF-11A) as well as cytokines and cytokine receptors involved in Th2 inflammation (IL-4RA, IL-6), NK cell regulation (IL18R1, IL15RA, IL27) and neutrophil recruitment (CXCL8/IL-8). Cytokine and interleukin signaling pathways were among the top ranked pathways associated with death (**Table 2**).

## Discussion

This study provides an assessment of the plasma proteome in a cohort of individuals with biopsy-confirmed kidney diseases. We evaluated associations of 225 plasma biomarkers with histopathologic findings in native kidney biopsies and subsequent risks of kidney disease progression and death and identified several new promising biomarker candidates. Pathways enriched in the plasma proteome of patients with kidney diseases point toward key mechanisms involved in disease pathogenesis, including inflammation, extracellular matrix remodeling, disturbances of hemostasis, cell growth, and apoptosis.

Previous large-scale proteomics studies have primarily focused on evaluation of the urine CKD proteome.^15-19^ Fewer studies have utilized proteomics to interrogate plasma in patients with CKD.^9, 20-22^ Some of the individual proteins that we investigated have previously been shown to be associated with histopathologic lesions and adverse clinical outcomes in individuals with kidney disease. Among those are plasma KIM-1^1, 23^ and fibroblast growth factor-23 (FGF-23).^4^ Many others are new findings that represent areas for future investigation.

### Biomarkers of Histopathology

In line with results from previous studies, KIM-1 was the biomarker most strongly associated with more severe ATI.^1, 2, 23^ VSIG2, a protein of unknown function, was the top-performing marker for IFTA. VSIG2 has been associated with incident heart failure^24^ and prevalent diabetes in the general population,^25^ but there are no studies, to our knowledge, that have linked VSIG2 to kidney diseases. Several inflammatory markers were associated with histopathologic lesions. EN-RAGE, a pro-inflammatory marker which leads to IL-1β and TNF-α release, had the strongest association with glomerular inflammation. In rodent models of CKD, EN-RAGE was reported to play a key role in the pathogenesis of vascular calcification through modulation of Pi co-transporter (Pit-1) expression.^26^ This finding is supported by previous studies in humans that described associations between elevated plasma EN-RAGE levels and atherosclerosis as well as cardiovascular and all-cause mortality in patients on dialysis.^27-29^ Our study adds that EN-RAGE may be crucial not only for sustaining chronic systemic inflammation in individuals with kidney disease but may also be an important marker specific for glomerular inflammation. The single marker that remained significantly associated with vascular pathology after multivariable adjustment was FABP2, a protein expressed in epithelial cells of the small intestines and involved in the reabsorption and distribution of long-chain fatty acids.^30^ Polymorphisms in the FABP2 gene have been linked to insulin resistance, dyslipidemia, and cardiovascular diseases.^30, 31^ We are not aware of any prior studies that have evaluated plasma FABP2 in kidney disease.

### Biomarkers of Adverse Clinical Events

A previous plasma proteomics study by Carlsson et al. of individuals from the general population identified 20 plasma proteins that were associated with incident CKD.^20^ Although the associations in the Carlsson study lost statistical significance after adjustment for baseline eGFR, seven of these proteins (TRAIL-R2, PGF, CD40, PAR-1, FGF-23, CSF-1, and TM) were also associated with kidney disease progression in our study and remained statistically significant after multivariable adjustment. We observed a particularly strong association for PGF, a member of the vascular endothelial growth factor (VEGF) family that has previously been studied in the setting of preeclampsia where lower circulating maternal PGF levels were found to be associated with the disease.^32, 33^ A prior study in kidney disease demonstrated that higher PGF levels associated with lower risks of AKI and mortality in patients undergoing cardiac surgery,^34^ but another study identified that higher PGF levels associated with an increased risk of incident CKD.^20^ We observed an increased risk of kidney disease progression with higher PGF levels. It is possible that in patients with established CKD, PGF levels may not have a pathogenic role but rather reflect a compensatory response to sustained angiogenic imbalance that is present in the setting of chronic injury. We also observed a strong association between higher levels of BAMBI and greater risk of kidney disease progression. BAMBI serves as a regulator of angiogenesis and endothelial homeostasis through modulation of alternative TGF-β signaling pathways such as extracellular signal-related kinase (ERK)1/2 and Smad1/5.^35, 36^ Diabetic BAMBI^−/−^ mice develop more severe albuminuria with increased activation of alternative TGF-β pathways compared to wildtype animals.^35^ In this study, we observed an association between higher, not lower, BAMBI levels and greater risks of kidney disease progression. Since our cohort primarily included patients with established CKD, these findings may point toward an important role of BAMBI in regulating anti-fibrotic responses while its elevation could be due to the active and ongoing fibrotic processes in CKD. This, however, remains to be tested in additional cohort studies of CKD.

The top-performing biomarker associated with increased risks of death was TRAIL-R2, a key mediator of apoptosis, which was also associated with kidney disease progression. TRAIL-R2 is a death domain-containing receptor that is capable of transmitting apoptotic signals in response to TRAIL binding.^37^ TRAIL, a member of the TNF gene superfamily, has been shown to associate with albuminuria in diabetes and plays an important role in the progression of diabetic nephropathy by inducing renal cell death.^20, 37^

We found lower levels of 5 plasma proteins to be associated with an increased risk of death. Among those was SCF, a regulator of bone marrow-derived stem cell migration and survival, known to act on early hematopoietic stem cells and primitive hematopoietic progenitor cells. Findings from rodent models suggest that SCF is crucial to mediate tubular epithelial cell survival after hypoxic injury.^38^ Similar findings were reported in mice in which local injection of SCF improved cardiac function after myocardial infarction.^39^ In the Malmo Diet and Cancer Study, lower circulating levels of SCF associated with significantly higher risks of stroke and heart failure as well as cardiovascular and all-cause mortality.^40^ We observed similar findings in the current study and found lower SCF levels to be associated with an increased risk of death in patients with kidney diseases. Evaluation of inflammatory parameters in the Malmoe Diet and Cancer Study also revealed that C-reactive protein levels and white blood cell counts correlated inversely with SCF.^40^ In line with this, we showed that SCF was inversely associated with both ATI and interstitial inflammation which could suggest that SCF may attenuate inflammation in patients with kidney disease or that in turn inflammation may function as a negative regulator of SCF. We also found that lower levels of ADAMTS13 were associated with a greater risk of death. ADAMTS13 is a von Willebrand factor (VWF) cleaving protease, and deficiency of plasma ADAMTS13 activity causes thrombotic thrombocytopenic purpura. In a case-control study of patients with malignant hypertension, ADAMTS13 levels correlated inversely with creatinine and were significantly decreased in patients with more severe thrombotic microangiopathy and hypertension.^41^ While the functional relevance and putative prognostic value of these proteins need to be further explored, several biomarkers shared common biologic pathways with pathophysiological relevance for kidney diseases, including inflammation and extracellular matrix remodeling, apoptosis, angiogenesis, and endothelial dysfunction.

### Strengths and Limitations

Significant strengths of our study include the large number of protein biomarkers included in the analyses. Adjudicated histopathologic scores on lesion severity and the prospective study design with long-term follow up data allowed us to test associations not only with histopathology but also with major adverse clinical events including kidney disease progression and death. Our study has several limitations that warrant consideration as well. First, our approach was exploratory, and we cannot determine causality. We were not able to take serum creatinine values prior to biopsy into account and did not account for therapy at baseline which could alter levels of biomarkers or an individual’s risk of disease progression or death. We were also not able to compare plasma protein measurements to protein measurements in the urine. Limitations of the proteomics assay include that protein values are obtained in relative quantification units (normalized protein expression values) rather than as absolute values which may limit comparability between studies. As we carefully selected 3 Olink proteomics panels based on their potential relevance for kidney disease, the selection of proteins included in the study limited our ability to perform unbiased pathway analyses.

In summary, we identified a number of protein biomarkers associated with histopathologic lesions on native kidney biopsy specimens and subsequent kidney disease progression and death. Several of our biomarker findings merit development of quantitative assays for further replication in other prospective cohort studies to develop markers for non-invasive diagnosis and optimal prognosis. Studies using techniques such as Mendelian randomization may identify which of our biomarker findings represent therapeutic targets for patients with specific kidney diseases. Our study demonstrates how findings from a cohort of individuals with available kidney pathology and prospective clinical follow-up can uncover relevant pathways and biomarker candidates for understanding, diagnosing, and providing prognosis of kidney diseases.

## Data Availability

Data from the Boston Kidney Biopsy Cohort are stored at the Brigham and Womens Hospital in Boston, MA. Data can be accessed upon request to the study PI Dr. Sushrut S. Waikar.

## Acknowledgements

We thank the members of the laboratory of S.S.W. for their invaluable assistance in the Boston Kidney Biopsy Cohort. I.M.S. is supported by the American Philosophical Society Daland Fellowship in Clinical Investigation. S.S.W. is also supported by NIH grants UH3DK114915, U01DK085660, U01DK104308, R01DK103784, and R21DK119751. A.S. is supported by NIH grant K23DK120811, NIDDK Kidney Precision Medicine Project Opportunity Pool grant under U2CDK114886, and core resources from the George M. O’Brien Kidney Research Center at Northwestern University (NU-GoKIDNEY) P30DK114857. M.Z. is supported by a NIH NIDDK T32 award DK007199. This work was conducted with support from Harvard Catalyst. The Harvard Clinical and Translational Science Center (National Center for Advancing Translational Sciences, National Institutes of Health Award UL1TR001102) and financial contributions from Harvard University and its affiliated academic healthcare centers. The content is solely the responsibility of the authors and does not necessarily represent the official views of Harvard Catalyst, Harvard University and its affiliated academic healthcare centers, or the NIH. Part of this work was presented as an oral presentation at the 2020 American Society of Nephrology Scientific Session on October 22^th^.

## Author Contributions

I.M.S, S.S.M, R.P., A.S., P.C.W., B.D.H., and S.S.W were responsible for the concept and design of the study. Z.A.K., A.A., M.Z., A.S., R.P. and S.S.W. adjudicated clinical outcomes. I.E.S. and H.G.R. were responsible for the adjudication of histopathology. S.S.M, I.M.S., and S.S.W. were responsible for statistical analyses. All authors interpreted the data. I.M.S. and S.S.W. drafted the manuscript. All authors contributed to critical revisions of the manuscript for important intellectual content.

