## Supplemental Material for "Circulating Plasma Biomarkers in Biopsy-Confirmed Kidney Disease: Results from the Boston Kidney Biopsy Cohort"

**Supplemental Table 1.** Histopathologic scoring system for light microscopy.

**Supplemental Table 2.** Associations of plasma biomarkers with histopathologic lesions.

**Supplemental Table 3.** Associations of plasma biomarkers with kidney disease progression.

**Supplemental Table 4.** Associations of plasma biomarkers with death.

**Supplemental Table 5.** Abbreviations and list of all plasma biomarkers included in the analyses.

**Supplemental Table 1.** Histopathologic scoring system for light microscopy

| Histologic Feature* | Scoring |
|---|---|
| Mesangial Matrix Expansion | 0 (none), 1 (mild,), 2 (moderate), 3 (severe) |
| Global Glomerulosclerosis | 0 (≤10%), 1 (11-25%), 2 (26-50%), 3 (>50%) |
| Segmental Glomerulosclerosis | 0 (≤10%), 1 (11-25%), 2 (26-50%), 3 (>50%) |
| Endocapillary Glomerular Inflammation | 0 (≤10%), 1 (11-25%), 2 (26-50%), 3 (>50%) |
| Extracapillary Cellular Crescents | 0 (≤10%), 1 (11-25%), 2 (26-50%), 3 (>50%) |
| Focal Glomerular Necrosis | 0 (≤10%), 1 (11-25%), 2 (26-50%), 3 (>50%) |
| Fibrocellular Crescents | 0 (≤10%), 1 (11-25%), 2 (26-50%), 3 (>50%) |
| Interstitial Fibrosis and Tubular Atrophy | 0 (≤10%), 1 (11-25%), 2 (26-50%), 3 (>50%) |
| Inflammation, Non-Fibrosed Interstitium | 0 (≤10%), 1 (11-25%), 2 (26-50%), 3 (>50%) |
| Inflammation, Fibrosed Interstitium | 0 (≤10%), 1 (11-25%), 2 (26-50%), 3 (>50%) |
| Acute Tubular Injury | 0 (none), 1 (mild,), 2 (moderate), 3 (severe) |
| Arterial Sclerosis | 0 (none), 1 (mild,), 2 (moderate), 3 (severe) |
| Arteriolar Sclerosis | 0 (none), 1 (mild,), 2 (moderate), 3 (severe) |

Percentages were calculated by assessing affected areas over total cortical volume or glomeruli affected.
References used to develop scoring system:

Mauer, S, Steffes, M, Ellis, E, Sutherland, D, Brown, D, Goetz, F: Structural-Functional Relationships in Diabetic Nephropathy. *J Clin Invest,* 74**:** 1143-1155, 1984.

Bajema, I, ECH, H, Hansen, B, Hermans, J, Noel, L, Waldherr, R, et al.: The renal histopathology in systemic vasculitis: an international survey study of inter- and intra-observer agreement. *Nephrol Dial Transplant,* 11**:** 1989-1995, 1996.

Racusen, LC, Solez, K, Colvin, RB, Bonsib, SM, Castro, MC, Cavallo, T, et al.: The Banff 97 working classification of renal allograft pathology. *Kidney Int,* 55**:** 713-723, 1999.

Weening, J, D'Agati, V, Schwartz, M, Seshan, S, Alpers, C, Appel, G, et al.: The Classification of Glomerulonephritis in Systemic Lupus Erythematosus Revisited. *Journal of the American Society of Nephrology,* 15**:** 241-250, 2004

D'Agati, VD, Fogo, AB, Bruijn, JA, Jennette, JC: Pathologic classification of focal segmental glomerulosclerosis: a working proposal. *American Journal of Kidney Diseases,* 43**:** 368-382, 2004.

Working Group of the International IgA Nephropathy Network and the Renal Pathology Society: Roberts, IS, Cook, HT, Troyanov, S, Alpers, CE, Amore, A, Barratt, J, et al.: The Oxford classification of IgA nephropathy: pathology definitions, correlations, and reproducibility. *Kidney Int,* 76**:** 546-556, 2009.

Tervaert, TW, Mooyaart, AL, Amann, K, Cohen, AH, Cook, HT, Drachenberg, CB, et al.: Pathologic classification of diabetic nephropathy. *J Am Soc Nephrol,* 21**:** 556-563, 2010.

Berden, AE, Ferrario, F, Hagen, EC, Jayne, DR, Jennette, JC, Joh, K, et al.: Histopathologic classification of ANCA-associated glomerulonephritis. *J Am Soc Nephrol,* 21**:** 1628-1636, 2010.

Farris, AB, Adams, CD, Brousaides, N, Della Pelle, PA, Collins, AB, Moradi, E, et al.: Morphometric and visual evaluation of fibrosis in renal biopsies. *J Am Soc Nephrol,* 22**:** 176-186, 2011.

**Supplemental Table 2.** Associations of plasma biomarkers with histopathologic lesions.

| Biomarker | % difference* | 95% CI | P value |
|---|---|---|---|
| **GLOMERULAR INFLAMMATION** | | | |
| **EN-RAGE** | 98.3 | 61.2, 144.0 | 6.28E-08 |
| **DNER** | -15.2 | -20.0, -10.1 | 1.16E-05 |
| **MCP-3** | 51.3 | 29.6, 76.5 | 5.34E-05 |
| **NOS3** | 45.5 | 23.5, 71.3 | 0.0021 |
| **Gal-9** | 19.0 | 10.2, 28.4 | 0.0025 |
| **IL-1ra** | 35.3 | 17.9, 55.2 | 0.0046 |
| **CSF-1** | 10.5 | 5.2, 16.1 | 0.0192 |
| **IL6** | 47.5 | 21.1, 79.5 | 0.0285 |
| **GDF-2** | -22.7 | -32.2, -11.9 | 0.0317 |
| **CCL4** | 32.0 | 14.5, 52.1 | 0.0325 |
| **hOSCAR** | 8.6 | 4.1, 13.3 | 0.0325 |
| **DPP6** | -13.8 | -20.3, -6.9 | 0.0424 |
| **CXCL10** | 52.7 | 22.5, 90.3 | 0.0435 |
| **PDCD1** | 15.5 | 7.1, 24.5 | 0.0484 |
| **INFLAMMATION, NON-FIBROSED INTERSTITIUM** | | | |
| **SCF** | -22.2 | -30.5, -13.0 | 0.0034 |
| **CXCL9** | 71.7 | 33.8, 120.3 | 0.0060 |
| **MMP7** | 21.7 | 11.1, 33.3 | 0.0065 |
| **DPP6** | -17.0 | -24.3, -9.0 | 0.0187 |
| **CCL23** | 23.9 | 11.0, 38.3 | 0.0362 |
| **CALCA** | 54.9 | 23.3, 94.5 | 0.0431 |
| **GLOMERULAR SCLEROSIS** | | | |
| **TF** | 17.2 | 10.7, 24.1 | 1.71E-05 |
| **PRSS27** | 23.7 | 14.4, 33.7 | 3.22E-05 |
| **SCF** | 22.5 | 13.5, 32.2 | 5.91E-05 |
| **GIF** | 43.2 | 24.6, 64.6 | 0.0001 |
| **CA14** | 18.0 | 9.1, 27.7 | 0.0102 |
| **PTX3** | -16.7 | -23.8, -9.0 | 0.0138 |
| **GDF-2** | 22.5 | 10.9, 35.4 | 0.0174 |
| **VSIG2** | 20.6 | 9.9, 32.4 | 0.0214 |
| **IL10** | -19.8 | -28.4, -10.1 | 0.0397 |
| **NPPC** | 25.0 | 11.3, 40.5 | 0.0433 |
| **MESANGIAL EXPANSION** | | | |
| **TRAIL** | 19.7 | 10.3, 29.9 | 0.0045 |
| **Gal-9** | 21.8 | 11.0, 33.7 | 0.0084 |
| **CSF-1** | 13.3 | 6.8, 20.3 | 0.0107 |
| **ENTPD6** | 13.6 | 6.8, 20.8 | 0.0150 |
| **PD-L2** | 18.7 | 9.0, 29.2 | 0.0219 |
| **TRANCE** | 42.1 | 19.0, 69.7 | 0.0268 |
| **ADM** | 24.2 | 11.3, 38.6 | 0.0290 |
| **CCL11** | 24.2 | 11.0, 39.0 | 0.0422 |
| **INTERSTITIAL FIBROSIS/TUBULAR ATROPHY** | | | |
| **VSIG2** | 30.0 | 15.7, 46.1 | 0.0029 |

| | | | |
|---|---|---|---|
| **TF** | 16.8 | 8.5, 25.7 | 0.0097 |
| **AMBP** | 10.1 | 5.1, 15.3 | 0.0134 |
| **IL-10RB** | 13.3 | 6.6, 20.4 | 0.0148 |
| **PGF** | 18.9 | 9.2, 29.4 | 0.017 |
| **FGF-23** | 47.4 | 21.7, 78.5 | 0.0194 |
| **CLEC1A** | 22.9 | 10.8, 36.3 | 0.0242 |
| **PTK7** | 22.1 | 10.3, 35.1 | 0.0296 |
| **PAR-1** | 16.8 | 7.9, 26.5 | 0.0340 |
| **TNFRSF9** | 32.8 | 14.5, 53.9 | 0.0441 |
| **INFLAMMATION, FIBROSED INTERSTITIUM** | | | |
| **MMP7** | 20.8 | 10.1, 32.5 | 0.0165 |
| **SPON2** | 8.4 | 4.1, 12.9 | 0.0250 |
| **KIM1** | 65.2 | 27.7, 113.7 | 0.0350 |
| **ARTERIOLAR SCLEROSIS** | | | |
| **FABP2** | 34.6 | 16.3, 55.9 | 0.0184 |
| **ACUTE TUBULAR INJURY** | | | |
| **SCF** | -29.5 | -37.7, -20.2 | 1.22E-05 |
| **KIM1** | 101.1 | 51.8, 166.4 | 0.0004 |
| **TGF-alpha** | 27.4 | 14.3, 42.0 | 0.0037 |
| **PLXDC1** | -15.8 | -22.3, -8.8 | 0.0066 |
| **PTX3** | 32.6 | 14.5, 53.6 | 0.0435 |

[*]Percent differences were derived from linear regression models using biomarkers as the outcome and each histopathologic lesion as the predictor variable. Models were further adjusted for age, sex, race, and eGFR. Percent differences in plasma biomarkers were calculated by raising 2 to the power of the beta-coefficient, subtracting 1, and multiplying by 100 [$(2^{\beta} - 1) * 100$)] for each respective histopathologic lesion. Shown are significant associations after applying Bonferroni correction. Name and abbreviation of each biomarker are shown in Supplemental Table 5.

Reference categories:
Reference is absence of lesion for glomerular inflammation and inflammation in the non-fibrosed interstitium;
Reference is none/mild lesion severity for mesangial expansion, acute tubular injury, and arteriolar sclerosis;
Reference is 0-25% of cortical volume affected for global glomerulosclerosis, inflammation in the fibrosed interstitium, and interstitial fibrosis/tubular atrophy.

**Supplemental Table 3.** Associations of plasma biomarkers with kidney disease progression.

| Biomarker | HR | 95% CI | P value |
|---|---|---|---|
| **PGF** | 5.4 | 3.4, 8.7 | 5.65E-10 |
| **BAMBI** | 3.0 | 2.1, 4.2 | 3.51E-07 |
| **TNFRSF11A** | 2.6 | 1.9, 3.5 | 3.68E-07 |
| **TRAIL-R2** | 2.8 | 2.0, 3.9 | 7.03E-07 |
| **CX3CL1** | 2.6 | 1.9, 3.6 | 3.61E-06 |
| **FGF-23** | 1.7 | 1.4, 2.0 | 3.64E-06 |
| **IL-15RA** | 6.0 | 3.2, 11.2 | 4.28E-06 |
| **PTK7** | 2.8 | 1.9, 3.9 | 5.13E-06 |
| **CA12** | 2.8 | 1.9, 4.2 | 7.64E-05 |
| **ADM** | 3.4 | 2.1, 5.6 | 0.0001 |
| **VEGFA** | 3.7 | 2.2, 6.1 | 0.0002 |
| **NUCB2** | 2.5 | 1.7, 3.6 | 0.0002 |
| **CD40** | 2.8 | 1.8, 4.2 | 0.0003 |
| **CSF-1** | 9.4 | 3.7, 23.7 | 0.0005 |
| **SPON2** | 32.1 | 7.5, 137.3 | 0.0007 |
| **KIM1** | 1.6 | 1.3, 1.9 | 0.0011 |
| **IL-4RA** | 2.1 | 1.5, 3.0 | 0.0019 |
| **DCN** | 3.9 | 2.1, 7.0 | 0.0023 |
| **CLEC1A** | 2.4 | 1.6, 3.5 | 0.0025 |
| **VSIG2** | 2.0 | 1.5, 2.7 | 0.0028 |
| **PAR-1** | 3.7 | 2.0, 6.8 | 0.0067 |
| **TNFRSF9** | 1.7 | 1.3, 2.2 | 0.0075 |
| **PD-L1** | 2.4 | 1.6, 3.6 | 0.0084 |
| **TNFRSF10A** | 2.3 | 1.5, 3.4 | 0.0120 |
| **TM** | 3.4 | 1.9, 6.1 | 0.0123 |
| **CAPG** | 1.8 | 1.4, 2.4 | 0.0149 |
| **IL-10RB** | 3.9 | 2.0, 7.9 | 0.0261 |
| **ENAH** | 2.2 | 1.5, 3.4 | 0.0306 |
| **IL16** | 1.8 | 1.3, 2.4 | 0.0322 |
| **TF** | 2.6 | 1.6, 4.4 | 0.0485 |

Shown are significant associations after applying Bonferroni correction. Models are adjusted for age, sex, race, log(proteinuria), eGFR, and primary clinicopathologic diagnosis. Name and abbreviation of each biomarker are shown in Supplemental Table 5. HR; Hazard Ratio

**Supplemental Table 4.** Associations of plasma biomarkers with death.

| Biomarker | HR | 95% CI | P value |
|---|---|---|---|
| **SCF** | 0.4 | 0.2, 0.5 | 7.43E-06 |
| **DSG4** | 0.5 | 0.4, 0.6 | 1.70E-05 |
| **CA14** | 0.4 | 0.3, 0.6 | 0.0040 |
| **PLXDC1** | 0.2 | 0.1, 0.4 | 0.0069 |
| **ADAM-TS13** | 0.1 | 0.0, 0.3 | 0.0274 |
| **TRAIL-R2** | 2.9 | 2.0, 4.0 | 2.79E-07 |
| **CDCP1** | 2.4 | 1.8, 3.3 | 4.31E-06 |
| **TNFRSF10A** | 2.7 | 1.9, 3.9 | 9.20E-06 |
| **PD-L1** | 2.4 | 1.7, 3.3 | 2.67E-05 |
| **BNP** | 1.3 | 1.2, 1.5 | 5.58E-05 |
| **CTSL1** | 3.0 | 2.0, 4.5 | 5.63E-05 |
| **PTK7** | 2.5 | 1.8, 3.6 | 7.93E-05 |
| **IL-4RA** | 2.3 | 1.7, 3.3 | 8.19E-05 |
| **IL8** | 1.6 | 1.3, 2.0 | 0.0002 |
| **ENTPD2** | 2.4 | 1.7, 3.3 | 0.0002 |
| **THBS2** | 8.2 | 3.5, 19.6 | 0.0004 |
| **ACE2** | 1.9 | 1.5, 2.5 | 0.0005 |
| **STX8** | 2.2 | 1.6, 3.1 | 0.0006 |
| **TOP2B** | 1.6 | 1.3, 1.9 | 0.0010 |
| **ENAH** | 2.6 | 1.7, 4.0 | 0.0011 |
| **IL6** | 1.5 | 1.3, 1.8 | 0.0013 |
| **IL-27** | 3.0 | 1.9, 5.0 | 0.0019 |
| **NOS3** | 1.6 | 1.3, 1.9 | 0.0022 |
| **Gal-9** | 4.9 | 2.4, 10.1 | 0.0024 |
| **ADM** | 3.7 | 2.0, 6.6 | 0.0034 |
| **OPG** | 2.7 | 1.7, 4.3 | 0.0046 |
| **IL-18R1** | 2.6 | 1.7, 4.0 | 0.0053 |
| **TNFRSF11A** | 1.9 | 1.4, 2.6 | 0.0068 |
| **CD40** | 2.6 | 1.6, 4.1 | 0.0093 |
| **AGRP** | 2.3 | 1.5, 3.4 | 0.0099 |
| **DCN** | 3.2 | 1.8, 5.7 | 0.0100 |
| **PGF** | 2.8 | 1.7, 4.6 | 0.0179 |
| **PRELP** | 9.5 | 3.1, 29.7 | 0.0220 |
| **CD4** | 2.9 | 1.7, 4.8 | 0.0236 |
| **IL-15RA** | 3.4 | 1.8, 6.5 | 0.0316 |

Shown are significant associations after applying Bonferroni correction. Models are adjusted for age, sex, race, log(proteinuria), eGFR, and primary clinicopathologic diagnosis. Name and abbreviation of each biomarker are shown in Supplemental Table 5. HR; Hazard Ratio.

**Supplemental Table 5.** Abbreviations and list of all plasma protein biomarkers included in the analyses.

| Abbreviation | Biomarker | Uniprot ID | Panel |
|---|---|---|---|
| ACE2 | Angiotensin-converting enzyme 2 (ACE2) | Q9BYF1 | CARDIOVASCULAR |
| ADA | Adenosine Deaminase (ADA) | P00813 | INFLAMMATION |
| ADAM-TS13 | A disintegrin and metalloproteinase with thrombospondin motifs 13 (ADAM-TS13) | Q76LX8 | CARDIOVASCULAR |
| ADGRG1 | Adhesion G-protein coupled receptor G1 (ADGRG1) | Q9Y653 | ORGAN DAMAGE |
| ADM | ADM (ADM) | P35318 | CARDIOVASCULAR |
| AGRP | Agouti-related protein (AGRP) | O00253 | CARDIOVASCULAR |
| AIFM1 | Apoptosis-inducing factor 1, mitochondrial (AIFM1) | O95831 | ORGAN DAMAGE |
| ALDH3A1 | Aldehyde dehydrogenase, dimeric NADP-preferring (ALDH3A1) | P30838 | ORGAN DAMAGE |
| AMBP | Protein AMBP (AMBP) | P02760 | CARDIOVASCULAR |
| AMN | Protein amnionless (AMN) | Q9BXJ7 | ORGAN DAMAGE |
| ANG-1 | Angiopoietin-1 (ANGPT1) | Q15389 | CARDIOVASCULAR |
| ATP6AP2 | Renin receptor (ATP6AP2) | O75787 | ORGAN DAMAGE |
| AXIN1 | Axin-1 (AXIN1) | O15169 | INFLAMMATION |
| BAMBI | BMP and activin membrane-bound inhibitor homolog (BAMBI) | Q13145 | ORGAN DAMAGE |
| BANK1 | B-cell scaffold protein with ankyrin repeats (BANK1) | Q8NDB2 | ORGAN DAMAGE |
| Beta-NGF | Beta-nerve growth factor (Beta-NGF) | P01138 | INFLAMMATION |
| BMP-6 | Bone morphogenetic protein 6 (BMP-6) | P22004 | CARDIOVASCULAR |
| BNP | Natriuretic peptides B (BNP) | P16860 | CARDIOVASCULAR |
| BOC | Brother of CDO (Protein BOC) | Q9BWV1 | CARDIOVASCULAR |
| BTC | Probetacellulin (BTC) | P35070 | ORGAN DAMAGE |
| CA12 | Carbonic anhydrase 12 (CA12) | O43570 | ORGAN DAMAGE |
| CA14 | Carbonic anhydrase 14 (CA14) | Q9ULX7 | ORGAN DAMAGE |
| CA5A | Carbonic anhydrase 5A, mitochondrial (CA5A) | P35218 | CARDIOVASCULAR |
| CALCA | Calcitonin (CALCA) | P01258 | ORGAN DAMAGE |
| CALR | Calreticulin (CALR) | P27797 | ORGAN DAMAGE |
| CAPG | Macrophage-capping protein (CAPG) | P40121 | ORGAN DAMAGE |
| CASP-8 | Caspase-8 (CASP-8 ) | Q14790 | INFLAMMATION |
| CCL11 | Eotaxin (CCL11) | P51671 | INFLAMMATION |
| CCL17 | C-C motif chemokine 17 (CCL17) | Q92583 | CARDIOVASCULAR |
| CCL19 | C-C motif chemokine 19 (CCL19) | Q99731 | INFLAMMATION |
| CCL20 | C-C motif chemokine 20 (CCL20) | P78556 | INFLAMMATION |

| | | | |
|---|---|---|---|
| CCL23 | C-C motif chemokine 23 (CCL23) | P55773 | INFLAMMATION |
| CCL25 | C-C motif chemokine 25 (CCL25) | O15444 | INFLAMMATION |
| CCL28 | C-C motif chemokine 28 (CCL28) | Q9NRJ3 | INFLAMMATION |
| CCL3 | C-C motif chemokine 3 (CCL3) | P10147 | INFLAMMATION |
| CCL4 | C-C motif chemokine 4 (CCL4 ) | P13236 | INFLAMMATION |
| CD244 | Natural killer cell receptor 2B4 (CD244) | Q9BZW8 | INFLAMMATION |
| CD4 | T-cell surface glycoprotein CD4 (CD4) | P01730 | CARDIOVASCULAR |
| CD40 | CD40L receptor (CD40) | P25942 | INFLAMMATION |
| CD40-L | CD40 ligand (CD40-L) | P29965 | CARDIOVASCULAR |
| CD5 | T-cell surface glycoprotein CD5 (CD5) | P06127 | INFLAMMATION |
| CD6 | T-cell differentiation antigen CD6 (CD6) | P30203 | INFLAMMATION |
| CD84 | SLAM family member 5 (CD84) | Q9UIB8 | CARDIOVASCULAR |
| CD8A | T-cell surface glycoprotein CD8 alpha chain (CD8A) | P01732 | INFLAMMATION |
| CDCP1 | CUB domain-containing protein 1 (CDCP1) | Q9H5V8 | INFLAMMATION |
| CEACAM8 | Carcinoembryonic antigenrelated cell adhesion molecule 8 (CEACAM8) | P31997 | CARDIOVASCULAR |
| CLEC1A | C-type lectin domain family 1 member A (CLEC1A) | Q8NC01 | ORGAN DAMAGE |
| CNTN2 | Contactin-2 (CNTN2) | Q02246 | ORGAN DAMAGE |
| CRH | Corticoliberin (CRH) | P06850 | ORGAN DAMAGE |
| CSF-1 | Macrophage colony-stimulating factor 1 (CSF-1) | P09603 | INFLAMMATION |
| CST5 | Cystatin D (CST5) | P28325 | INFLAMMATION |
| CTRC | Chymotrypsin C (CTRC) | Q99895 | CARDIOVASCULAR |
| CTSL1 | Cathepsin L1 (CTSL1) | P07711 | CARDIOVASCULAR |
| CX3CL1 | Fractalkine (CX3CL1 ) | P78423 | INFLAMMATION |
| CXCL1 | C-X-C motif chemokine 1 (CXCL1) | P09341 | INFLAMMATION |
| CXCL10 | C-X-C motif chemokine 10 (CXCL10 ) | P02778 | INFLAMMATION |
| CXCL11 | C-X-C motif chemokine 11 (CXCL11) | O14625 | INFLAMMATION |
| CXCL5 | C-X-C motif chemokine 5 (CXCL5 ) | P42830 | INFLAMMATION |
| CXCL6 | C-X-C motif chemokine 6 (CXCL6) | P80162 | INFLAMMATION |
| CXCL9 | C-X-C motif chemokine 9 (CXCL9 ) | Q07325 | INFLAMMATION |
| DCN | Decorin (DCN) | P07585 | CARDIOVASCULAR |
| DECR1 | 2,4-dienoyl-CoA reductase, mitochondrial (DECR1) | Q16698 | CARDIOVASCULAR |
| Dkk-1 | Dickkopf-related protein 1 (Dkk-1) | O94907 | CARDIOVASCULAR |
| DNER | Delta and Notch-like epidermal growth factor-related receptor (DNER) | Q8NFT8 | INFLAMMATION |
| DPP6 | Dipeptidyl aminopeptidase-like protein 6 (DPP6) | P42658 | ORGAN DAMAGE |

| | | | |
|---|---|---|---|
| DSG4 | Desmoglein-4 (DSG4) | Q86SJ6 | ORGAN DAMAGE |
| EGFL7 | Epidermal growth factor-like protein 7 (EGFL7) | Q9UHF1 | ORGAN DAMAGE |
| EN-RAGE | Protein S100-A12 (EN-RAGE ) | P80511 | INFLAMMATION |
| ENAH | Protein enabled homolog (ENAH) | Q8N8S7 | ORGAN DAMAGE |
| ENTPD2 | Ectonucleoside triphosphate diphosphohydrolase 2 (ENTPD2) | Q9Y5L3 | ORGAN DAMAGE |
| ENTPD6 | Ectonucleoside triphosphate diphosphohydrolase 6 (ENTPD6) | O75354 | ORGAN DAMAGE |
| EPO | Erythropoietin (EPO) | P01588 | ORGAN DAMAGE |
| FABP2 | Fatty acid-binding protein, intestinal (FABP2) | P12104 | CARDIOVASCULAR |
| FABP9 | Fatty acid-binding protein 9 (FABP9) | Q0Z7S8 | ORGAN DAMAGE |
| FGF-19 | Fibroblast growth factor 19 (FGF-19) | O95750 | INFLAMMATION |
| FGF-21 | Fibroblast growth factor 21 (FGF-21) | Q9NSA1 | CARDIOVASCULAR |
| FGF-23 | Fibroblast growth factor 23 (FGF-23) | Q9GZV9 | CARDIOVASCULAR |
| FGF-5 | Fibroblast growth factor 5 (FGF-5) | P12034 | INFLAMMATION |
| FGR | Tyrosine-protein kinase Fgr (FGR) | P09769 | ORGAN DAMAGE |
| Flt3L | Fms-related tyrosine kinase 3 ligand (Flt3L) | P49771 | INFLAMMATION |
| FOSB | Protein fosB (FOSB) | P53539 | ORGAN DAMAGE |
| FOXO1 | Forkhead box protein O1 (FOXO1) | Q12778 | ORGAN DAMAGE |
| FS | Follistatin (FS) | P19883 | CARDIOVASCULAR |
| Gal-9 | Galectin-9 (Gal-9) | O00182 | CARDIOVASCULAR |
| GALNT10 | Polypeptide N-acetylgalactosaminyltransferase 10 (GALNT10) | Q86SR1 | ORGAN DAMAGE |
| GDF-2 | Growth/differentiation factor 2 (GDF-2) | Q9UK05 | CARDIOVASCULAR |
| GDNF | Glial cell line-derived neurotrophic factor (GDNF) | P39905 | INFLAMMATION |
| GH | Growth hormone (GH) | P01241 | CARDIOVASCULAR |
| GIF | Gastric intrinsic factor (GIF) | P27352 | CARDIOVASCULAR |
| GLO1 | Lactoylglutathione lyase (GLO1) | Q04760 | CARDIOVASCULAR |
| GT | Gastrotropin (GT) | P51161 | CARDIOVASCULAR |
| HAOX1 | Hydroxyacid oxidase 1 (HAOX1) | Q9UJM8 | CARDIOVASCULAR |
| HB-EGF | Proheparin-binding EGF-like growth factor (HB-EGF) | Q99075 | CARDIOVASCULAR |
| HGF | Hepatocyte growth factor (HGF) | P14210 | INFLAMMATION |
| HO-1 | Heme oxygenase 1 (HO-1) | P09601 | CARDIOVASCULAR |
| hOSCAR | Osteoclast-associated immunoglobulin-like receptor (hOSCAR) | Q8IYS5 | CARDIOVASCULAR |
| HPGDS | Hematopoietic prostaglandin D synthase (HPGDS) | O60760 | ORGAN DAMAGE |
| HSP 27 | Heat shock 27 kDa protein (HSP 27) | P04792 | CARDIOVASCULAR |
| IDUA | Alpha-L-iduronidase (IDUA) | P35475 | CARDIOVASCULAR |

| | | | |
|---|---|---|---|
| IgG Fc Rec IIb | Low affinity immunoglobulin gamma Fc region receptor II-b (IgG Fc receptor II-b) | P31994 | CARDIOVASCULAR |
| IL-10RA | Interleukin-10 receptor subunit alpha (IL-10RA) | Q13651 | INFLAMMATION |
| IL-10RB | Interleukin-10 receptor subunit beta (IL-10RB) | Q08334 | INFLAMMATION |
| IL-12B | Interleukin-12 (IL-12) | P29460 | INFLAMMATION |
| IL-15RA | Interleukin-15 receptor subunit alpha (IL-15RA) | Q13261 | INFLAMMATION |
| IL-17A | Interleukin-17A (IL-17A) | Q16552 | INFLAMMATION |
| IL-17C | Interleukin-17C (IL-17C) | Q9P0M4 | INFLAMMATION |
| IL-17D | Interleukin-17D (IL-17D) | Q8TAD2 | CARDIOVASCULAR |
| IL-18R1 | Interleukin-18 receptor 1 (IL-18R1) | Q13478 | INFLAMMATION |
| IL-1ra | Interleukin-1 receptor antagonist protein (IL-1ra) | P18510 | CARDIOVASCULAR |
| IL-20RA | Interleukin-20 receptor subunit alpha (IL-20RA) | Q9UHF4 | INFLAMMATION |
| IL-27 | Interleukin-27 (IL-27) | Q8NEV9 | CARDIOVASCULAR |
| IL-2RB | Interleukin-2 receptor subunit beta (IL-2RB) | P14784 | INFLAMMATION |
| IL-4RA | Interleukin-4 receptor subunit alpha (IL-4RA) | P24394 | CARDIOVASCULAR |
| IL10 | Interleukin-10 (IL10) | P22301 | INFLAMMATION |
| IL16 | Pro-interleukin-16 (IL16) | Q14005 | CARDIOVASCULAR |
| IL18 | Interleukin-18 (IL-18) | Q14116 | INFLAMMATION |
| IL1RL2 | Interleukin-1 receptor-like 2 (IL1RL2) | Q9HB29 | CARDIOVASCULAR |
| IL6 | Interleukin-6 (IL6) | P05231 | INFLAMMATION |
| IL7 | Interleukin-7 (IL-7) | P13232 | INFLAMMATION |
| IL8 | Interleukin-8 (IL-8) | P10145 | INFLAMMATION |
| INPPL1 | Phosphatidylinositol 3,4,5-trisphosphate 5-phosphatase 2 (INPPL1) | O15357 | ORGAN DAMAGE |
| ITGB1BP2 | Melusin (ITGB1BP2) | Q9UKP3 | CARDIOVASCULAR |
| KIM1 | Kidney Injury Molecule 1 (KIM1) | Q96D42 | ORGAN DAMAGE |
| KIR3DL1 | Killer cell immunoglobulin-like receptor 3DL1 (KIR3DL1) | P43629 | ORGAN DAMAGE |
| LAP TGF-beta-1 | Latency-associated peptide transforming growth factor beta-1 (LAP TGF-beta-1) | P01137 | INFLAMMATION |
| LAT2 | Linker for activation of T-cells family member 2 (LAT2) | Q9GZY6 | ORGAN DAMAGE |
| LEP | Leptin (LEP) | P41159 | CARDIOVASCULAR |
| LHB | Lutropin subunit beta (LHB) | P01229 | ORGAN DAMAGE |
| LIF-R | Leukemia inhibitory factor receptor (LIF-R) | P42702 | INFLAMMATION |
| LOX-1 | Lectin-like oxidized LDL receptor 1 (LOX-1) | P78380 | CARDIOVASCULAR |
| LPL | Lipoprotein lipase (LPL) | P06858 | CARDIOVASCULAR |
| LRP1 | Prolow-density lipoprotein receptor-related protein 1 (LRP1) | Q07954 | ORGAN DAMAGE |
| MAGED1 | Melanoma-associated antigen D1 (MAGED1) | Q9Y5V3 | ORGAN DAMAGE |

| | | | |
|---|---|---|---|
| MAP4K5 | Mitogen-activated protein kinase kinase kinase kinase 5 (MAP4K5) | Q9Y4K4 | ORGAN DAMAGE |
| MARCO | Macrophage receptor MARCO (MARCO) | Q9UEW3 | CARDIOVASCULAR |
| MCP-1 | Monocyte chemotactic protein 1 (MCP-1) | P13500 | INFLAMMATION |
| MCP-2 | Monocyte chemotactic protein 2 (MCP-2) | P80075 | INFLAMMATION |
| MCP-3 | Monocyte chemotactic protein 3 (MCP-3) | P80098 | INFLAMMATION |
| MCP-4 | Monocyte chemotactic protein 4 (MCP-4) | Q99616 | INFLAMMATION |
| MERTK | Tyrosine-protein kinase Mer (MERTK) | Q12866 | CARDIOVASCULAR |
| MMP-1 | Matrix metalloproteinase-1 (MMP-1) | P03956 | INFLAMMATION |
| MMP-10 | Matrix metalloproteinase-10 (MMP-10) | P09238 | INFLAMMATION |
| MMP12 | Matrix metalloproteinase-12 (MMP-12) | P39900 | CARDIOVASCULAR |
| MMP7 | Matrix metalloproteinase-7 (MMP-7) | P09237 | CARDIOVASCULAR |
| MVK | Mevalonate kinase (MVK) | Q03426 | ORGAN DAMAGE |
| NBN | Nibrin (NBN) | O60934 | ORGAN DAMAGE |
| NCF2 | Neutrophil cytosol factor 2 (NCF2) | P19878 | ORGAN DAMAGE |
| NEMO | NF-kappa-B essential modulator (NEMO) | Q9Y6K9 | CARDIOVASCULAR |
| NOS3 | Nitric oxide synthase, endothelial (NOS3) | P29474 | ORGAN DAMAGE |
| NPPC | C-type natriuretic peptide (NPPC) | P23582 | ORGAN DAMAGE |
| NT-3 | Neurotrophin-3 (NT-3) | P20783 | INFLAMMATION |
| NUCB2 | Nucleobindin-2 (NUCB2) | P80303 | ORGAN DAMAGE |
| OPG | Osteoprotegerin (OPG) | O00300 | INFLAMMATION |
| OSM | Oncostatin-M (OSM) | P13725 | INFLAMMATION |
| PAPPA | Pappalysin-1 (PAPPA) | Q13219 | CARDIOVASCULAR |
| PAR-1 | Proteinase-activated receptor 1 (PAR-1) | P25116 | CARDIOVASCULAR |
| PARP-1 | Poly (ADP-ribose) polymerase 1 (PARP-1) | P09874 | CARDIOVASCULAR |
| PD-L1 | Programmed cell death 1 ligand 1 (PD-L1) | Q9NZQ7 | INFLAMMATION |
| PD-L2 | Programmed cell death 1 ligand 2 (PD-L2) | Q9BQ51 | CARDIOVASCULAR |
| PDCD1 | Programmed cell death protein 1 (PDCD1) | Q15116 | ORGAN DAMAGE |
| PDGF subunit B | Platelet-derived growth factor subunit B (PDGF subunit B) | P01127 | CARDIOVASCULAR |
| PDGFC | Platelet-derived growth factor C (PDGFC) | Q9NRA1 | ORGAN DAMAGE |
| PGF | Placenta growth factor (PGF) | P49763 | ORGAN DAMAGE |
| PIgR | Polymeric immunoglobulin receptor (PIgR) | P01833 | CARDIOVASCULAR |
| PLIN1 | Perilipin-1 (PLIN1) | O60240 | ORGAN DAMAGE |
| PLXDC1 | Plexin domain-containing protein 1 (PLXDC1) | Q8IUK5 | ORGAN DAMAGE |
| PON2 | Serum paraoxonase/arylesterase 2 (PON2) | Q15165 | ORGAN DAMAGE |

| | | | |
|---|---|---|---|
| PRELP | Prolargin (PRELP) | P51888 | CARDIOVASCULAR |
| PRKAB1 | 5'-AMP-activated protein kinase subunit beta-1 (PRKAB1) | Q9Y478 | ORGAN DAMAGE |
| PRSS27 | Serine protease 27 (PRSS27) | Q9BQR3 | CARDIOVASCULAR |
| PRSS8 | Prostasin (PRSS8 ) | Q16651 | CARDIOVASCULAR |
| PSGL-1 | P-selectin glycoprotein ligand 1 (PSGL-1) | Q14242 | CARDIOVASCULAR |
| PTK7 | Inactive tyrosine-protein kinase 7 (PTK7) | Q13308 | ORGAN DAMAGE |
| PTN | Pleiotrophin (PTN) | P21246 | ORGAN DAMAGE |
| PTX3 | Pentraxin-related protein PTX3 (PTX3) | P26022 | CARDIOVASCULAR |
| PVALB | Parvalbumin alpha (PVALB) | P20472 | ORGAN DAMAGE |
| PXN | Paxillin (PXN) | P49023 | ORGAN DAMAGE |
| RAGE | Receptor for advanced glycosylation end products (RAGE) | Q15109 | CARDIOVASCULAR |
| RARRES1 | Retinoic acid receptor responder protein 1 (RARRES1) | P49788 | ORGAN DAMAGE |
| RASSF2 | Ras association domain-containing protein 2 (RASSF2) | P50749 | ORGAN DAMAGE |
| REN | Renin (REN) | P00797 | CARDIOVASCULAR |
| RRM2B | Ribonucleoside-diphosphate reductase subunit M2 B (RRM2B) | Q7LG56 | ORGAN DAMAGE |
| SCF/c-KIT ligand | Stem cell factor (SCF) | P21583 | INFLAMMATION |
| SERPINA12 | Serpin A12 (SERPINA12) | Q8IW75 | CARDIOVASCULAR |
| SERPINA9 | Serpin A9 (SERPINA9) | Q86WD7 | ORGAN DAMAGE |
| SIRT2 | SIR2-like protein 2 (SIRT2) | Q8IXJ6 | INFLAMMATION |
| SLAMF1 | Signaling lymphocytic activation molecule (SLAMF1) | Q13291 | INFLAMMATION |
| SLAMF7 | SLAM family member 7 (SLAMF7) | Q9NQ25 | CARDIOVASCULAR |
| SOD2 | Superoxide dismutase (Mn), mitochondrial (SOD2) | P04179 | CARDIOVASCULAR |
| SORT1 | Sortilin (SORT1) | Q99523 | CARDIOVASCULAR |
| SPON2 | Spondin-2 (SPON2) | Q9BUD6 | CARDIOVASCULAR |
| SRC | Proto-oncogene tyrosine-protein kinase Src (SRC) | P12931 | CARDIOVASCULAR |
| ST1A1 | Sulfotransferase 1A1 (ST1A1) | P50225 | INFLAMMATION |
| ST3GAL1 | CMP-N-acetylneuraminate-beta-galactosamide-alpha-2,3-sialyltransferase 1 (ST3GAL1) | Q11201 | ORGAN DAMAGE |
| STAMBP | STAM-binding protein (STAMPB) | O95630 | INFLAMMATION |
| STK4 | Serine/threonine-protein kinase 4 (STK4) | Q13043 | CARDIOVASCULAR |
| STX8 | Syntaxin-8 (STX8) | Q9UNK0 | ORGAN DAMAGE |
| TF/ F3 | Tissue factor (TF) | P13726 | CARDIOVASCULAR |
| TGF-alpha | Transforming growth factor alpha (TGF-alpha) | P01135 | INFLAMMATION |
| TGM2 | Protein-glutamine gamma-glutamyltransferase 2 (TGM2) | P21980 | CARDIOVASCULAR |
| THBS2 | Thrombospondin-2 (THBS2) | P35442 | CARDIOVASCULAR |

| | | | |
|---|---|---|---|
| THPO | Thrombopoietin (THPO) | P40225 | CARDIOVASCULAR |
| TIE2 | Angiopoietin-1 receptor (TIE2) | Q02763 | CARDIOVASCULAR |
| TIGAR | Fructose-2,6-bisphosphatase TIGAR (TIGAR) | Q9NQ88 | ORGAN DAMAGE |
| TM | Thrombomodulin TM | P07204 | CARDIOVASCULAR |
| TMPRSS15 | Enteropeptidase (TMPRSS15) | P98073 | ORGAN DAMAGE |
| TNFB | TNF-beta (TNFB) | P01374 | INFLAMMATION |
| TNFRSF10A | Tumor necrosis factor receptor superfamily member 10A (TNFRSF10A) | O00220 | CARDIOVASCULAR |
| TNFRSF11A | Tumor necrosis factor receptor superfamily member 11A (TNFRSF11A) | Q9Y6Q6 | CARDIOVASCULAR |
| TNFRSF13B | Tumor necrosis factor receptor superfamily member 13B (TNFRSF13B) | O14836 | CARDIOVASCULAR |
| TNFRSF9 | Tumor necrosis factor receptor superfamily member 9 (TNFRSF9) | Q07011 | INFLAMMATION |
| TNFSF14 | Tumor necrosis factor ligand superfamily member 14 (TNFSF14 ) | O43557 | INFLAMMATION |
| TNNI3 | Troponin I, cardiac muscle (TNNI3) | P19429 | ORGAN DAMAGE |
| TOP2B | DNA topoisomerase 2-beta (TOP2B) | Q02880 | ORGAN DAMAGE |
| TRAIL/TNFSF10 | TNF-related apoptosis-inducing ligand (TRAIL) | P50591 | INFLAMMATION |
| TRAIL-R2/ TNFRSF10B | TNF-related apoptosis-inducing ligand receptor 2 (TRAIL-R2) | O14763 | CARDIOVASCULAR |
| TRANCE | TNF-related activation-induced cytokine (TRANCE) | O14788 | INFLAMMATION |
| TWEAK | Tumor necrosis factor (Ligand) superfamily, member 12 (TWEAK) | O43508 | INFLAMMATION |
| uPA | Urokinase-type plasminogen activator (uPA) | P00749 | INFLAMMATION |
| VEGFA | Vascular endothelial growth factor A (VEGF-A) | P15692 | INFLAMMATION |
| VEGFD | Vascular endothelial growth factor D (VEGFD) | O43915 | CARDIOVASCULAR |
| VSIG2 | V-set and immunoglobulin domain-containing protein 2 (VSIG2) | Q96IQ7 | CARDIOVASCULAR |
| XCL1 | Lymphotactin (XCL1) | P47992 | CARDIOVASCULAR |
| YES1 | Tyrosine-protein kinase Yes (YES1) | P07947 | ORGAN DAMAGE |
| 4E-BP1 | Eukaryotic translation initiation factor 4E-binding protein 1 (4E-BP1) | Q13541 | INFLAMMATION |
